# Estimating the burden of leptospirosis in the Caribbean: Insights from environmental and sociodemographic factors

**DOI:** 10.64898/2025.12.19.25342643

**Authors:** Beatris Mario Martin, Zhonghan Zhang, Holly Jian, Sebastian Vernal, Eric J. Nilles, Luis Furuya-Kanamori, Benn Sartorius, Colleen L Lau

**Affiliations:** University of Queensland Centre for Clinical Research (UQCCR), Faculty of Health, Medicine, and Behavioural Sciences, The University of Queensland, Brisbane, Australia; Harvard Humanitarian Initiative, Cambridge, Massachusetts, USA; Harvard Medical School, Boston, Massachusetts, USA; Brigham and Women’s Hospital, Boston, Massachusetts, USA

**Author notes:** **Corresponding author**: Beatris Mario Martin.

## Abstract

**Introduction:** Previous studies up to early 2000s found that leptospirosis’ incidence in humans was high across the Caribbean region (CR), yet up-to-date and reliable surveillance data are scarce. Limited research capacity in the region has further contributed to less robust characterisation of transmission drivers, perpetuating a cycle of neglect. To address these gaps and support evidence-based public health responses, this study aims to update incidence estimates in the CR by integrating data from multiple data sources, including peer-reviewed publications, surveillance reports and environmental and sociodemographic datasets.

**Methodology/Principal Findings:** We used mixed-effects hierarchical Poisson models at the country/territory-year level, incorporating covariates i.e., precipitation, temperature, gross domestic product, biodiversity loss, human footprint, water flow accumulation index, populations exposed to forested areas and frequency of extreme weather events to estimate annual case numbers by country/territory. Temporal patterns in case fatality rate (CFR) were modelled using locally estimated scatterplot smoothing regressions. Between 2001 and 2023, we estimated 32,311 (95% CI 28,646-36,832) cases in the region. Annual incidence (cases/100,000 population) showed a decrease trend, with small- to medium-sized population islands exhibiting the highest predicted incidence (Guadeloupe 22.7 (95%CI 20.5-25.0) and Saint Vincent and the Grenadines 22.3 (95%CI 17.6-27.9)). Estimated CFR increased over the study period, from 8.8% (95%CI 4.1-13.4) (2001) to 12.2% (.2.2-22.2) (2022).

**Conclusions/Significance:** Leptospirosis remains an important and often overlooked public health concern in the CR, where small island developing states bear a disproportionate burden of the disease. These findings underscore the urgent need for strengthening and integrating the surveillance systems and laboratory capacity in the region, particularly in small states, to provide accurate data to support cost-effective public health interventions.

**Authors summary:** Leptospirosis is a bacterial infection that affects both animals and humans and is common across the Caribbean. Yet, many countries and territories in the region lack reliable and up-to-date information on disease incidence and mortality. This is partly due to limited laboratory capacity, inconsistent case reporting, and competing health priorities. As a result, the true burden of leptospirosis remains unclear, especially for resource-limited small island nations, which are vulnerable to potential impacts of climate change and extreme weather events on leptospirosis transmission.

In this study, we combined information from peer-reviewed publications, national surveillance reports, and environmental and sociodemographic datasets to estimate annual incidence and mortality across the Caribbean. Using statistical modelling, we estimated over 32,000 cases between 2001 and 2023, with small- and medium-sized islands showing the highest incidence (cases/100,000 population). We also estimated a slight increase in case-fatality rate over time.

Our findings highlight that small island developing states carry a disproportionate burden of this disease. Improving local surveillance systems and strengthening regional level collaboration are essential to help countries better identify risk, prepare for outbreaks, and reduce illness and deaths.

## INTRODUCTION

Leptospirosis is the most widespread bacterial zoonotic disease in the world (1) due to the *Leptospira*’s ability to infect a wide range of mammals and its potential to survive long periods in warm and humid environments including water and soil(2). Globally, 1 million cases/year and nearly 60,000 deaths are estimated(3). Across the Caribbean region (CR), previous studies up to early 2000s found that leptospirosis was highly endemic in humans, with countries like Guadeloupe, Martinique, Trinidad and Tobago, and Barbados presenting some of the highest annual reported incidence globally (ranging from 10.0 to 22.5 cases/ 100,000 population, respectively), only after Seychelles (Indian Ocean) (43.2/100,000 population) (4, 5). Additionally, between 1970 and 2012, one-third of all leptospirosis outbreaks occurred in the Latin America and Caribbean region (6), reflecting the region’s vulnerability to leptospirosis transmission (7).

Up to the early 1990s, leptospirosis was frequently associated with occupational risk and resource-poor rural environments (8–10). More recently, outbreaks have been increasingly associated with extreme weather events, such as heavy rainfall, tropical storms and hurricanes, which are frequently followed by floods (11, 12). As global temperatures rise, these events will become more severe (13), and can further represent a heavier burden to Small Island Developing States (SIDS), which are particularly vulnerable to the impact of climate change, both economically and environmentally (13). Additionally, SIDS are more vulnerable to biodiversity loss, as the reduction of terrestrial mammals can be associated with rodent population growth, which in turn is associated with increased leptospirosis transmission (14).

In the CR, environmental and sociodemographic conditions are ideal for *Leptospira* survival and transmission. First, the region has a very warm and humid climate, with high rainfall, particularly during the rainy season (from June to November) (1, 7, 15, 16). Second, rapid, unplanned urbanisation, poor sanitation in growing slums, and rising rodent population facilitate human contact with animal reservoirs and contaminated environments (12, 17), driving leptospirosis outbreaks in the CR, particularly following floods. Third, the traditional occupational risk remains an important source of infection, contributing to an endemic transmission associated with activities that require working in close contact with animals (e.g., dairy farmers, abattoir workers), or exposure to contaminated environments (e.g., sugarcane fieldworkers, sewerage workers) (18).

Despite its recognised burden, up-to-date and reliable leptospirosis data in the CR remain scarce (19). Efforts to characterise leptospirosis burden face persistent challenges, frequently resulting in potential underestimation of its true incidence. Leptospirosis surveillance is inconsistent across the CR, including variation in case definitions, irregular reporting, and limited access to laboratory diagnostics to confirm cases (20). These issues result in fragmented and outdated data, hindering effective public health planning and policy-making. The lack of robust data perpetuates a cycle of neglect, where insufficient attention and funding further limit research and surveillance. In resource-limited settings, public health systems often prioritise the most urgent health threats. In this context, shifting public health priorities can impact diagnosis and reporting capacity, particularly when multiple endemic diseases present as similar undifferentiated febrile syndromes. Recent events, such as the COVID-19 pandemic and the progressively larger dengue outbreaks in the CR, may have contributed to irregular, less robust, and poorly funded support for other endemic disease surveillance.

In the face of limited and less robust surveillance data, modelling offers a powerful tool to estimate disease burden (21), overcome data scarcity and guide decision-making. Predictive models are widely used in public health to assess epidemiological patterns and support health policy-making. In infectious disease epidemiology, statistical modelling can be used to investigate transmission dynamics, evaluate intervention impact, and identify vulnerable populations. By integrating diverse data sources, modelling can help overcome data gaps and provide actionable insights for decision-makers. To better understand the current impact of leptospirosis in the CR and support evidence-based public health responses, this study aims to provide updated incidence estimates by combining data from multiple sources, including peer-reviewed publications, surveillance reports and environmental and sociodemographic data.

## METHODS

### Geographic scope

In this study, we focused on 27 tropical SIDS in the CR. The particular interest in CR Islands Countries and Territories (CRICTs) was driven by higher leptospirosis transmission risk in tropical small island settings compared to continental coastal areas (3, 12, 14). The CRICTs included in this study were Anguilla, Antigua and Barbuda, Aruba, Bahamas, Barbados, British Virgin Islands, Cayman Islands, Cuba, Curaçao, Dominica, Dominican Republic, Grenada, Guadeloupe, Haiti, Jamaica, Martinique, Montserrat, Puerto Rico, Saint Barthélemy, Saint Kitts and Nevis, Saint Lucia, Saint Martin, Saint Vincent and the Grenadines, Sint Maarten, Trinidad and Tobago, Turks and Caicos, and U.S. Virgin Islands (**Figure 1**).

**Figure 1.**
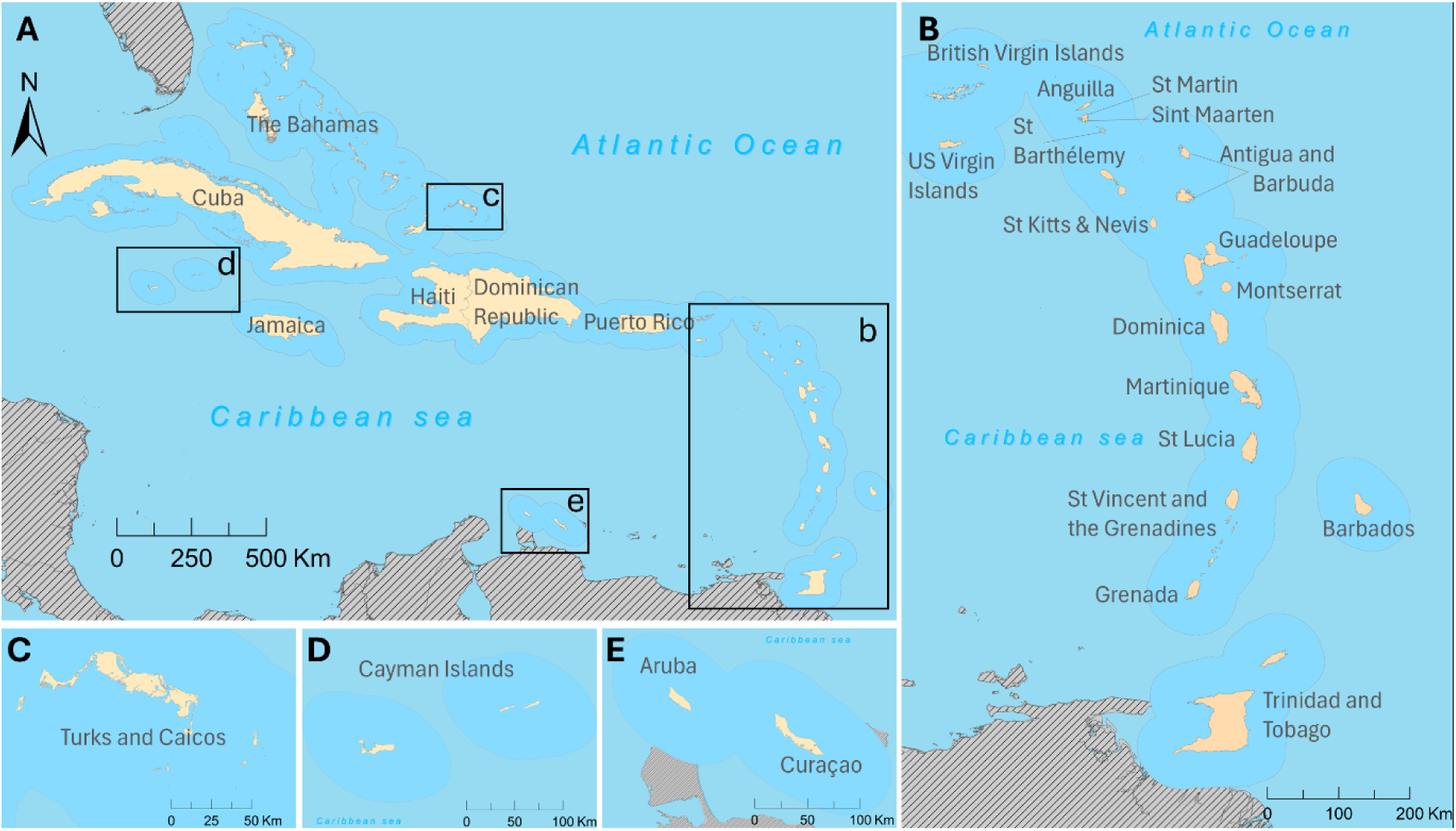
Caribbean island countries and territories included in the incidence model. A) Caribbean Region, B) Lesser Antilles group, and Trinidad and Tobago. C) Turks and Caicos, D) Cayman Islands, E) Aruba and Curaçao. Base layer from: https://datacatalog.worldbank.org/search/dataset/0038272/world-bank-official-boundaries (CC BY 4.0).

### Data Source and data extraction

#### Leptospirosis reported cases

To identify all relevant data for creating our incidence estimates model, we used three main data sources. First, we incorporated the total number of reported cases by year and location extracted from peer-reviewed publications included in a scoping review of leptospirosis morbidity and mortality in the CR between 2000 and 2023 (19). The complete list of peer-reviewed publications and their case definition are available in ***Supporting Table 1***. Second, we searched for grey literature on websites of health and other government departments and other relevant administrative levels to identify any epidemiological reports (e.g., weekly reports, epidemiological summaries and bulletins) communicating leptospirosis cases in a defined period. The full description of the grey literature search and included publications is available in ***Supporting Tables 2 and 3***. Finally, we incorporated reported cases available from the Pan American Health Organization (PAHO) Core Indicators Dashboard (22). When reports identified through grey literature did not include leptospirosis case definition, data (number of cases and/or incidence) were extracted as reported. We included confirmed cases from peer-reviewed publications and all reported cases from grey literature, assuming that reported cases should represent confirmed cases.

**Table 1.** Variables included in the mixed-effect model used to estimate leptospirosis incidence in the Caribbean region between 2000 and 2023, and the rationale for including each variable.

| Variable | Rationale for inclusion |
| --- | --- |
| Population density | Higher population density can increase exposure risk through closer contact with contaminated environments and limited sanitation infrastructure. Population density can be used as a proxy for urbanisation (42). |
| **Number of people exposed to each LCLU<sup>1</sup>** | Because the risk of leptospirosis varies across environments, and it is most relevant where people are exposed, estimating the number of people living within each LCLU class provides a more accurate measure of environment-specific transmission risk (43). |
| **Precipitation** | Heavy rainfall and flooding facilitate the spread of *Leptospira* bacteria by increasing waterborne transmission and environmental contamination (44, 45). It also contributes to displacement of rodent populations, increasing the geographic spread of the reservoir. |
| **Temperature** | Warmer temperatures can enhance bacterial survival in the environment, influencing seasonal patterns of leptospirosis outbreaks (46, 47). |
| **Biodiversity** | Reduced biodiversity may impact host-pathogen dynamics, potentially increasing the prevalence of reservoir species like rodents (14, 43). |
| **Human footprint** | Human activities such as urbanisation and land conversion can disrupt ecosystems and increase contact with contaminated environments (42). |
| **Water flow accumulation** | Areas with high water accumulation are more likely to retain contaminated water, increasing exposure risk during floods or heavy rains (48). |
| **GDP PPP<sup>2</sup>** | Lower economic development is often linked to inadequate sanitation, poor housing, and limited healthcare access, all of which elevate leptospirosis risk (49). |
| **Extreme weather events** | The CR<sup>3</sup> is frequently affected by extreme weather events, mainly heavy rainfall and hurricanes, often resulting in flooding. We extracted data (year, type of weather event, and number of events/year) from two regional climate and weather reports and from peer-reviewed publications to create a heatmap of 'water-related' extreme events (**Supporting Figure 1**) (35-41). |

<sup>1</sup>LCLU: land cover and land use. <sup>2</sup>GDP PPP: gross domestic product based on purchasing power parity. <sup>3</sup>CR: Caribbean Region.

The number of leptospirosis cases by CRICTs and year was extracted and recorded in a Microsoft Excel spreadsheet (Microsoft Corporation, Redmond, Washington, USA) (23). For CRICTs and years in which leptospirosis cases were reported in more than one data source and there was disagreement between them, peer-reviewed studies were considered the primary source, followed by government reports. Finally, the numbers of reported cases available at the PAHO Core Indicators Dashboard were only used if no other data source was available for that CRICT and year. The final number of cases by year and CRICT used in our model can be found in ***Supporting Table 4***.

#### Mortality data

Data on mortality and case fatality rate (CFR) were obtained from two sources. First, using the same search strategy used to identify publications reporting leptospirosis cases, we expanded the timeframe of the peer-reviewed studies (1979-2022) by removing any date restriction. The decision to expand the time frame was based on the limited number of peer-reviewed studies reporting mortality data identified by the scoping review. The full list of peer-reviewed studies included in this step is listed in the ***Supporting Table 5***. Second, we included all the epidemiological reports reporting mortality, identified through grey literature search described above. Mortality data extracted from both sources were combined, following the same hierarchy previously presented for cases. For each year, the number of reported deaths were combined to calculate CFR at the regional level and used to create a time trend.

#### Population data

Population data by CRICTs and year were obtained through the World Bank Group (24), and data from 1995 to 2023 were extracted. Missing population data were interpolated based on a linear regression model aggregated by year and CRICT.

#### Sociodemographic and environmental variables

Variables included in the model aimed to reflect the complex drivers of leptospirosis transmission (7) (Table 1). We extracted population density (25), number of people exposed to each land cover/land use (26), precipitation (27), temperature (28), biodiversity loss(29), human footprint (30), water flow accumulation(31), gross domestic product purchasing power parity (GDP PPP) (32–34), the annual frequency each country/territory was affected by a water related extreme weather event (heavy rainfall, storm, hurricane, flood) (35–41) and the country/territory’s total area in km^2^. Although extremely relevant, the availability of data on animal production (livestock distribution, type and size) were limited and could not be included in our analysis. A comprehensive description of all variables extracted and tested, the source from which the data were extracted, and how the data were processed before being included in the model is available in ***Supporting List 1*.**

In summary, for land cover/land use (LCLU), precipitation, temperature, and gross domestic product (GDP) data extracted were summarised, calculating the annual mean, between 2000 and 2023, for each CRICT. For biodiversity loss, water flow accumulation and human footprint, a unique value for each CRICT was used, and for extreme weather events, the sum of all water-related events from each year and CRICT was used. Extracted data were normalised using Z-score. Missing data were interpolated using linear models. Biodiversity loss data was unavailable for the Dutch territory of Sint Maarten, and data from the neighbouring territory of Saint Martin was used as a proxy.

### Morbidity and mortality model

The number of reported cases by location and year extracted from all incidence reports was used to create an overall incidence for the CR (*ε*), by dividing the sum of all reported cases (20,040) by the total population (573,031,662; considering only countries or territories and years for which there were observed cases) between 2001 and 2023. The overall incidence, *ε* (3.50), was incorporated into the final model as an offset. By including an offset into the model, we accounted for variations in observations across the study period and improved the model’s fit to the original data.

#### Variable selection and correlation analysis

Univariable Poisson mixed-effect regression models were developed to examine the association between leptospirosis reported cases (outcome variable) and the socioeconomic and environmental factors (covariates) and select potential variables to be included in the multivariable model. Covariates associated with leptospirosis reported cases and with a *P*-value below 0.1 were tested for correlation to be retained in the final model. Correlation (*ρ*) between environmental and sociodemographic covariates was assessed using Pearson correlation coefficients, and pairs of covariates with a *ρ* value ≥±0.6 were assessed by log-marginal likelihood. Among each pair of correlated covariates, we retained the covariate in the model with the highest log-marginal likelihood **(*Supporting* *Figure 2*)**.

**Figure 2.**
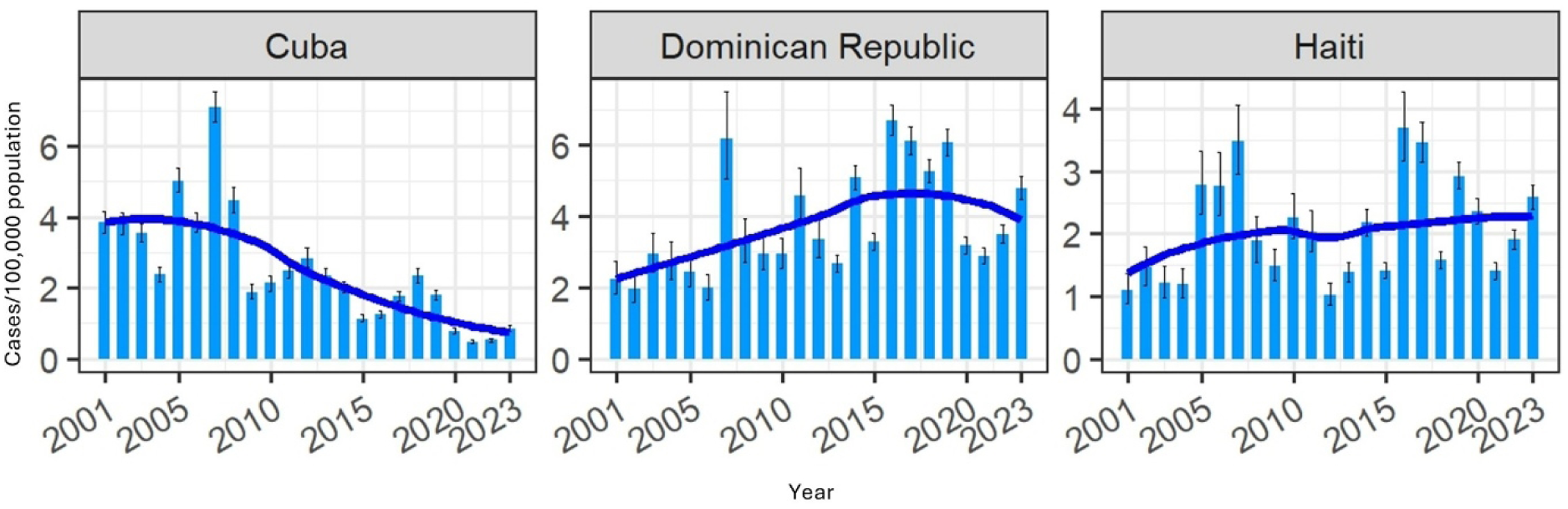
Estimated leptospirosis incidence by year in Cuba, Dominican Republic and Haiti, 2001-2023. The dark blue line represents an incidence time trend, using a locally estimated scatterplot smoothing regression (span 0.75).

#### Data analysis and prediction model

A multivariable, Poisson mixed-effect regression model was developed using the Integrated Nested Laplace Approximation (INLA) framework (50) to estimate leptospirosis incidence for each CRICT by year. CRICT and year were included as random effects (intercepts), modelled respectively as independent and identically distributed (iid) and as a first-order random walk (RW1) process. Penalised Complexity (PC) priors were assigned to the precision parameters of the random effects, implying that the standard deviation is greater than one with 1% probability. Sociodemographic and environmental variables were included as fixed effects. Weakly informative Gaussian priors (mean = 0, precision = 0.0001) were assigned for fixed effect coefficients. Model performance was evaluated using the root mean square error (RMSE) and cross-validation. The dataset was randomly divided into four subsets; three were used for model training and one for testing. This process was repeated ten times, and the RMSE and the Pearson correlation between observed and predicted cases were calculated for each iteration. The mean RMSE and correlation value were reported as the final performance metric.

Model adequacy and potential overdispersion were evaluated through posterior predictive checks. Using 500 posterior samples drawn from the fitted INLA model, replicated datasets were simulated under the fitted Poisson likelihood. Observed and simulated summary statistics (variance-to-mean ratio and proportion of zeros) were compared, and posterior predictive *P*-values were computed to assess model fit.

To calculate the predicted number of deaths by CRICT and year, we used the regional case fatality rate (CFR) trend. First, the annual CFR was calculated for years in which there were mortality data available. To do so, all reported deaths were divided by all reported cases (combining multiple countries when there were mortality data from more than one country/territory for that year). Annual data available were used to create a time trend plot. Then, using a Locally Estimated Scatterplot Smoothing (LOESS) model, we identified the fraction of observed data to be included in the smoothing process (span), and finally used these final parameters to build a LOESS model to predict CFR across 2001 to 2022 and the 95% CI. CFR was used to estimate deaths based on the results of the incidence model (number of deaths = predicted CFR multiplied by predicted incidence). The input data are available at **Supporting Table 6**.

R statistical programming language (R version 4.1.3, 2022-03-10) (51) was used to process spatial referenced environmental and sociodemographic data, statistical model to predict incidence was fitted using ‘*INLA’* package, for data visualisation ‘*ggplot2’* and ‘*ComplexHeatmap’* were used. Google Earth Engine (27) was used to access and extract precipitation and temperature data. Esri® ArcGIS PRO was used to create maps (52).

### Ethical considerations

All data used to build our model were obtained from publicly available sources, and no formal ethical approval was required.

## RESULTS

### Morbidity model

Between 2001 and 2023, it was estimated that 32,311 (95%CI 28,646-36,832) cases of leptospirosis occurred in the CR, with approximately 1,405 (95%CI 1,246-1,601) cases annually. Overall incidence for the study period was 3.37 (95%CI 2.99-3.85) cases/100,000 population (95%CI 3.13-3.99), showing a decreasing trend (**Table 2**). Predicted cases and incidence (cases/ 100,000 population) ranged from 755 (681-841) and 1.73 (1.56-1.92), in 2021, to 2,538 (2,237-2,897) cases and 6.27 (5.52-7.15), in 2007. Combined, the first decade (2001-2010) of the estimates (15,305) represented 47.4% of all cases in the study period (32,311) with an incidence of 3.82 (95%CI 3.33-4.46) cases/100,000 population.

**Table 2.** Leptospirosis annual reported cases, predicted cases, incidence and deaths for the Caribbean Region, between 2001 and 2023.

| Year | Observed/<br>reported<br>cases | Predicted Cases<br>(95% CI) | Predicted incidence per<br>100,000<br>(95%CI) | Predicted Deaths<sup>1</sup><br>(95%CI) |
| --- | --- | --- | --- | --- |
| 2001 | 620 | 1,405 (1,212-1,673) | 3.64 (3.14-4.34) | 123 (49-225) |
| 2002 | 640 | 1,325 (1,146-1,567) | 3.40 (2.94-4.02) | 118 (47-215) |
| 2003 | 781 | 1,640 (1,417-1,959) | 4.17 (3.61-4.99) | 148 (59-273) |
| 2004 | 514 | 1,242 (1,063-1,466) | 3.14 (2.68-3.70) | 113 (45-204) |
| 2005 | 969 | 1,712 (1,510-1,957) | 4.29 (3.79-4.91) | 155 (65-270) |
| 2006 | 855 | 1,515 (1,331-1,740) | 3.77 (3.31-4.33) | 136 (58-237) |
| 2007 | 1,332 | 2,538 (2,237-2,897) | 6.27 (5.52-7.15) | 227 (98-393) |
| 2008 | 662 | 1,569 (1,350-1,833) | 3.85 (3.31-4.49) | 141 (59-248) |
| 2009 | 325 | 1,028 (880-1,207) | 2.50 (2.14-2.94) | 93 (40-164) |
| 2010 | 577 | 1,330 (1,154-1,549) | 3.22 (2.79-3.75) | 122 (55-212) |
| 2011 | 611 | 1,892 (1,647-2,189) | 4.55 (3.96-5.27) | 177 (82-300) |
| 2012 | 424 | 1,060 (915-1,232) | 2.54 (2.19-2.95) | 101 (48-170) |
| 2013 | 667 | 1,114 (994-1,264) | 2.65 (2.36-3.00) | 108 (54-176) |
| 2014 | 1,319 | 1,667 (1,513-1,859) | 3.94 (3.58-4.39) | 164 (84-263) |
| 2015 | 880 | 888 (804-991) | 2.09 (1.89-2.33) | 89 (45-144) |
| 2016 | 1,133 | 1,587 (1,434-1,765) | 3.71 (3.36-4.13) | 163 (80-265) |
| 2017 | 1,273 | 1,804 (1,653-1,985) | 4.20 (3.85-4.63) | 190 (89-312) |
| 2018 | 1,284 | 1,417 (1,299-1,557) | 3.29 (3.02-3.62) | 154 (65-259) |
| 2019 | 1,475 | 1,592 (1,469-1,738) | 3.68 (3.40-4.02) | 177 (67-308) |
| 2020 | 817 | 931 (847-1,027) | 2.14 (1.94-2.36) | 107 (33-196) |
| 2021 | 716 | 755 (681-841) | 1.73 (1.56-1.92) | 89 (21-173) |
| 2022 | 1,028 | 1,060 (959-1,177) | 2.42 (2.19-2.68) | 129 (21-261) |
| 2023 | 1,139 | 1,239 (1,131-1,363) | 2.81 (2.57-3.10) | NA |

<sup>1</sup>Total number of predicted deaths per year. NA – not available.

The Dominican Republic (8,882; 7,937–9,930), Cuba (6,592; 6,079–7,138) and Haiti (4,838; 4,233–5,515) accounted for the largest absolute number of estimated cases, representing 63% of all cases in the region. However, they did not rank highest in incidence rate, with estimated rates (ranks) of 3.84 (9^th^), 2.55 (15^th^), and 2.09 (16^th^) cases per 100,000 population, respectively. In contrast, smaller islands such as Guadeloupe (2,124), St Vincent and the Grenadines (556 cases), Grenada (379), and Martinique (1,289) contributed only to 13% of total cases but had the four highest estimated incidences: 22.70 (20.53–25.04), 22.32 (17.60–27.92), 14.62 (11.82–17.90), and 14.53 (12.98–16.21) cases per 100,000 population, respectively **(Table 3)**.

**Table 3.** Mean annually predicted cases and incidence of leptospirosis by island countries and territories in the Caribbean Region, between 2001and 2023.

| Country | Observed/<br>reported<br>cases | Predicted cases<br>(mean, [95%CI]) | Predicted incidence<br>(mean, [95%CI]/100,000<br>population) |
| --- | --- | --- | --- |
| Anguilla | no data | 1 (0–6) | 0.40 (0.01–2.07) |
| Antigua and Barbuda | 10 | 56 (28–99) | 2.82 (1.43–5.02) |
| Aruba | no data | 1 (0–4) | 0.04 (0.00–0.16) |
| Bahamas | 19 | 22 (14–34) | 0.26 (0.16–0.39) |
| Barbados | 113 | 343 (276–422) | 5.42 (4.35–6.67) |
| British Virgin Islands | no data | 3 (0–13) | 0.41 (0.02–1.92) |
| Cayman Islands | 1 | 14 (2–49) | 1.09 (0.17–3.70) |
| Cuba | 6,589 | 6,592 (6,079–7,138) | 2.55 (2.35–2.76) |
| Curaçao | no data | 2 (0–9) | 0.07 (0.01–0.26) |
| Dominica | 127 | 127 (104–153) | 8.02 (6.57–9.70) |
| Dominican Republic | 5,321 | 8,882 (7,937–9,930) | 3.84 (3.43–4.29) |
| Grenada | 114 | 379 (307–465) | 14.62 (11.81–17.90) |
| Guadeloupe | 1,925 | 2,124 (1,921–2,342) | 22.70 (20.53–25.04) |
| Haiti | 1,590 | 4,838 (4,233–5,515) | 2.09 (1.83–2.38) |
| Jamaica | 1,057 | 2,277 (2,045–2,529) | 3.59 (3.22–3.99) |
| Martinique | 996 | 1,289 (1,152–1,438) | 14.53 (12.98–16.21) |
| Montserrat | no data | 1 (0–7) | 1.20 (0.03–6.49) |
| Puerto Rico | 1,100 | 3,329 (2,910–3,792) | 4.06 (3.55–4.63) |
| Saint Barthelemy | no data | 4 (0–25) | 1.87 (0.01–11.18) |
| Sint Maarten | no data | 2 (0–15) | 0.27 (0.00–1.82) |
| St. Kitts and Nevis | 23 | 29 (18–43) | 2.67 (1.72–3.96) |
| St. Lucia | 242 | 351 (299–409) | 8.89 (7.58–10.37) |
| St. Martin | no data | 6 (0–41) | 0.81 (0.00–5.20) |
| St. Vincent and the Grenadines | 89 | 556 (439–696) | 22.32 (17.60–27.92) |
| Trinidad and Tobago | 724 | 992 (882–1,111) | 3.22 (2.87–3.61) |
| Turks and Caicos Islands | 1 | 4 (1–12) | 0.50 (0.08–1.62) |
| US Virgin Islands | no data | 86 (0–537) | 3.47 (0.02–21.68) |

Over the 23-year study period, we did not identify any reports of leptospirosis cases in nine CRICTs (Anguilla, Aruba, British Virgin Islands, Curaçao, Montserrat, Saint Barthélemy, Sint Maarten, St. Martin, and US Virgin Islands). However, there is historical evidence of previous transmission and/or seroprevalence suggesting the occurrence of human leptospirosis in at least four of these nine CRICTs (i.e., Anguilla (53), British Virgin Islands (53), Montserrat (54), and US Virgin Islands (37)), hence, our model predicted outcomes for all 27 CRICTs, even those with no observed cases. For all nine CRICTs without leptospirosis observed case in the period investigated here, the predicted number of cases and incidence were negligible and unlikely to overestimate regional numbers.

### Temporal distribution by CRICT

In most CRICTs, there was a decreasing trend in the number of cases estimated during the study period. Cuba presented the greatest reduction in the total cases estimated, from 427 (394–463) in 2001 to 95 (85–105) in 2023, with an incidence drop from 3.83 (3.53–4.15) to 0.86 (0.77–0.96) cases/100,000 population. The Dominican Republic and Haiti were the only two countries in which the predicted number of cases increased over the years **(Figure 2)**. Graphs for all 27 CRICTs are presented in the ***Supporting* *Figure 3***.

**Figure 3.**
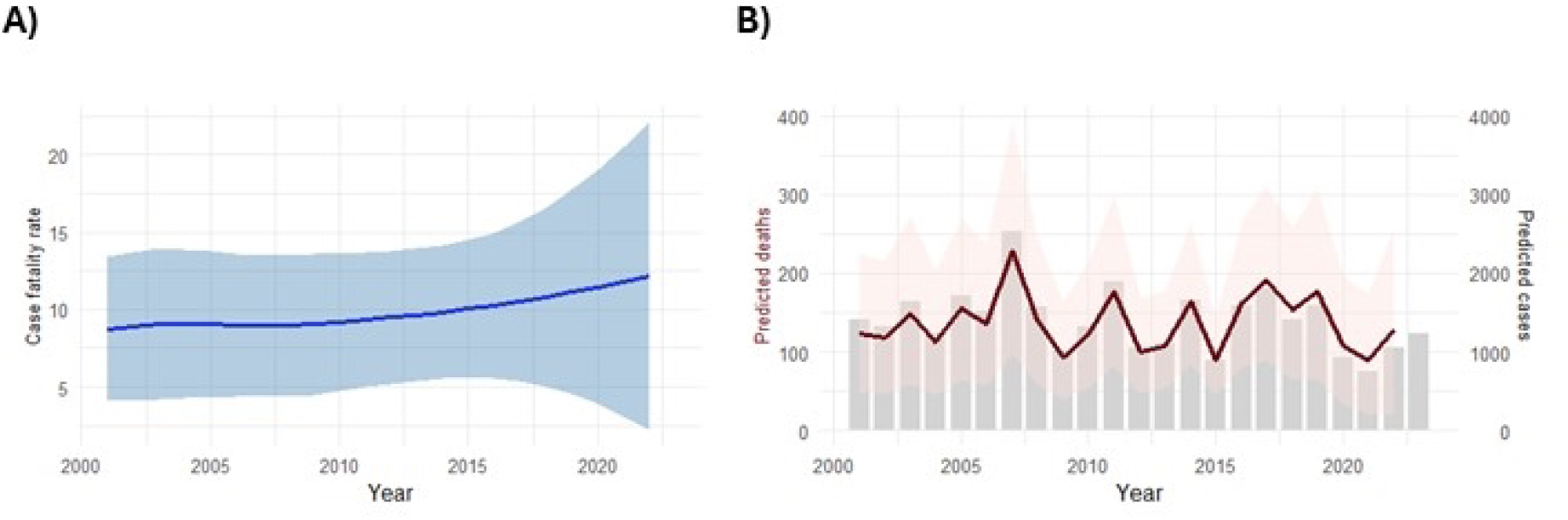
Calculated case fatality rate (CFR) and predicted deaths in the Caribbean region, 2001-2022. A) Smoothed CFR over time based on the locally estimated scatterplot smoothing regression. B) Predicted deaths are shown by the red line (left y-axis), with pink ribbon representing the 95% CI. Overall predicted cases are shown by the grey columns (right y-axis).

### Incidence model performance

The mean RSME was 115.04, corresponding to approximately 8% of the mean predicted value, and the mean Pearson correlation was 0.77 **(*Supporting Figure 4*)**, suggesting a high agreement between observed and predicted cases. Posterior predictive checks indicated that the Poisson mixed-effects model provided an adequate fit to the data. The observed and simulated variance-to-mean ratios were nearly identical (302.6 vs. 302.9), with a posterior predictive *P*-value of 0.5, suggesting no evidence of overdispersion. The proportion of zero counts in the observed data (33.2%) was slightly lower than that predicted by the model (37.9%), with a posterior predictive *P*-value of 0.97, indicating a mild tendency to overestimate zeros. Overall, the model captured the variability and distribution of the observed counts well, supporting the suitability of the Poisson likelihood for this analysis.

### Mortality estimates

Data extracted from 21 publications were used to create a CFR time trend for the CR between 1968 and 2022 (***Supporting Figure 5***). A LOESS curve was fitted using a span of 0.75, and the predicted CFR and 95%CI were extracted for the period between 2001 to 2022 (***Supporting Table 7***). CFR varied from 8.8% (95% CI 4.1-13.4%) in 2001 to 12.2% (95%CI 2.2-22.2%) in 2022, suggesting an increasing trend over the study period (**Figure 3a**). Using the predicted CFR, mortality was estimated based on the predicted incidence (**Figure 3b**). Overall, 3,025 (95%CI 1,263-5,266) deaths were predicted. Although peaks in mortality were mostly driven by peaks in incidence (e.g., 2007 is the year with the highest predicted incidence and mortality), the increasing trend impacted mortality peaks in years that were not among the highest incidence (e.g., 2019) (**Table 1**). There were not sufficient data from each CRICTs to model country/territory-level specific mortality.

## DISCUSSION

This is the first model, to our knowledge, to estimate annual leptospirosis incidence and mortality at the national level for the CR. By integrating multiple data sources and incorporating the frequency of water-related extreme weather events, our study was able to provide up-to-date estimates of leptospirosis morbidity and mortality data, address peaks in cases associated with these events and provide a better understanding of the disease’s epidemiological patterns in the CR. Our findings underscore the continued health burden caused by leptospirosis, estimating approximately 1,405 (95%CI 1,246-1,601) cases/100,000 population annually, despite the overall decline in morbidity over the study period and the scarce literature identified by our previous scoping review (19). Notably, our results suggest that small- to medium-population-size island countries and territories, particularly in the Lesser Antilles, exhibited disproportionately high incidence rates (e.g., St. Vincent and the Grenadines 22.32 cases/100,000 population), frequently exceeding the regional average (3.37 cases/100,000 population), highlighting the heterogeneous vulnerability across the region.

Additionally, while incidence declined in most CRICTs, it increased in the two countries on the Hispaniola island (Haiti and the Dominican Republic). By unravelling this national-level variation across the CR, our results reinforce the needs to improve local surveillance and to strengthening regional collaboration to support laboratory capacity and diagnoses.

The high incidence estimated for the small- to medium-population-size island countries and territories, such as St. Vincent and the Grenadines, Guadeloupe, Grenada, and Martinique reinforces the vulnerability of smaller, resource-limited CRICTs to leptospirosis transmission(14, 20). At the same time, these CRICTs were mostly underrepresented in our scoping review (19) compared to other medium- to low-incidence CRICTs (e.g., Jamaica and Puerto Rico). This important mismatch between estimated morbidity and documentation of burden demonstrates the importance of incidence models to support public health decision-making when limited data are available. For example, in the US Virgin Islands, our search did not identify any reported cases through peer-reviewed or grey literature in the last 23 years. However, the predicted incidence estimated by our model was not negligible, suggesting potential underreporting and/or environmental suitability for leptospirosis transmission. This is reinforced by a serosurvey conducted in the territory in 2019, which detected 4% seroprevalence despite no human cases documented before the Hurricanes Irma and Maria (37). The absence of reported cases likely reflects the neglected status of leptospirosis rather than its true absence. Moreover, including CRICTs such as Anguilla, Aruba, Curaçao, and Montserrat did not inflate regional estimates and instead provided valuable insights into environmental and sociodemographic suitability for transmission, thereby supporting proactive surveillance and prevention strategies.

The Dominican Republic, Cuba, and Haiti accounted for 63% of all estimated cases, largely due to their population size. However, Guadeloupe stands out as an exception—despite its relatively small population (<500,000 people), it ranks among the top in both case numbers (2,124) and incidence (22.70/100,000 population. This aligns with previous studies documenting the historical and recent impact of leptospirosis in Guadeloupe, including outbreaks and high seroprevalence (5, 55–57). Across the study period, Guadeloupe incidence showed only a small decrease, in contrast to other historical high-endemicity CRICTs, such as Jamaica, Trinidad and Tobago, Cuba and Barbados, where leptospirosis cases have been consistently reducing over the years (58), and in which our model estimated that recent incidence has been below the regional average.

However, the overall decreasing trend in leptospirosis incidence across the study period should be interpreted with caution. Although it could reflect the impact of public health interventions (e.g., animal vaccination and prophylaxis for people with temporary risk (6)) and broader societal improvements (e.g., flood prevention and management), it might be biased by data scarcity, which was more pronounced in the latest years of our study. The delay between investigation and publication might explain why there were fewer studies reporting leptospirosis data in more recent years. However, data scarcity was also observed among grey literature, becoming even more challenging during and after the COVID-19 pandemic period. Additionally, the occurrence of multiple and competing health crises (e.g., Zika, dengue, COVID-19) might have impacted shifts in disease prioritisation (59, 60). These concurrent challenges may have disrupted surveillance systems, strained healthcare access, overwhelmed systems and reduced diagnostic capacity, leading to underreporting, particularly of mild cases. At the same time, severe cases (e.g., those requiring hospitalisation and deaths) are less likely to be missed by surveillance systems, which may partially explain the slight increase in CFR more recently.

This study has several limitations and data gaps. Our incidence model was based on *Leptospira*’s life cycle and human transmission framework; while we created a robust dataset integrating multiple publicly available data sources to inform drivers and observed cases, it is important to acknowledge potential impacts of limited data availability. First, data on biodiversity loss were not available for the Dutch territory of Sint Maarten, requiring the use of proxy data from the neighbouring territory of Saint Martin. While this may reduce accuracy for that territory, the estimated cases were low and aligned with the absence of reported cases, suggesting minimal impact on overall model validity. Second, animal data (both livestock and rodent populations) were restricted to few countries/territories, thus not included into the model. As humans are an incidental host, and do not contribute to the transmission cycle, incorporating animal data would improve our model by capturing the impact of human exposure to animal reservoirs. Finally, the mortality model could not provide CRICTs-level estimates. The observed CFR varied across CRICTs and years and, unfortunately, data on healthcare access and quality were unavailable for most of the CRICTs during the study period. The lack of a reliable measure to adjust CFR based on access to healthcare hindered the possibility of providing a more comprehensive characterisation of leptospirosis mortality in the region. This study has also important strengths. We conducted a comprehensive literature review, comparing results from multiple data sources, to identify all reported cases and deaths associated to leptospirosis in the region. Additionally, by incorporating environmental and sociodemographic covariates in our estimates model, we were able to mitigate the limitations of data scarcity. Finally, we incorporated extreme water-related events to our model, allowing potential peaks in reported cases linked to those events to be addressed by the model.

Strengthening national health system data can contribute to improving data availability and reducing uncertainties regarding mortality rate across the region. Data gaps were particularly pronounced in smaller islands, which could further increase their vulnerability to leptospirosis ongoing transmission and outbreaks due lack of robust surveillance and research. As demonstrated by the US Virgin Islands, the absence of reports does not equate to the absence of transmission.

Leptospirosis remains an important and often overlooked public health concern in the CR, particularly in small island countries and territories. These same countries and territories often face data limitations, hindering the effort to fully characterise leptospirosis epidemiology and disease burden. This mismatch between vulnerability and data availability highlights the urgent need for more robust and coordinated regional surveillance systems. The CR could benefit from a regional integrated surveillance strategy. By adopting a unified case definition and strengthening laboratory capacity, disease monitoring can be improved by providing accurate data that can be used to guide targeted public health interventions and ensuring that leptospirosis receives appropriate attention within the regional health agenda.

## Acknowledgements

We thank Dr Eloise Skinner for support and dedication to help us find environmental georeferenced data to incorporate in our analysis. This work was supported by the Operational Research and Decision Support for Infectious Diseases (ODeSI) program, which is funded by The University of Queensland’s Health Research Accelerator (HERA) initiative (2021-2028).

## Data Availability Statement

All data analysed in the manuscript were obtained from publicly available sources, and source details are provided throughout the manuscript and supporting documents.

## Supporting Information

***Supporting Table 1.*** Peer-reviewed publications reporting leptospirosis in the Caribbean region, between 2001 and 2023, from which leptospirosis cases were extracted.

***Supporting Table 2.*** Grey literature search strategy used in Google Advanced to identify epidemiological reports of leptospirosis in the Caribbean region, between 2000 and 2023.

***Supporting Table 3.*** List of documents included through grey literature search by country/territory

***Supporting Table 4.*** Observed cases by country/territory and by year extracted from peer-reviewed studies, grey literature and Pan American Health Organization (PAHO) Core Indicators Dashboard.

***Supporting Table 5.*** Peer-reviewed publications used to identify the leptospirosis regional case-fatality rate

***Supporting List 1.*** A comprehensive list of variables assessed to be included in the mixed-effect model, their data sources and the spatial scale from the original data.

***Supporting Figure 1.*** Extreme weather events in the Caribbean Region between 2000 and 2023, by country and territory.

***Supporting Figure 2.*** Correlation Matrix showing the correlation between environmental and sociodemographic variables assessed using Pearson correlation coefficients. A ρ value ≥±0.6 indicates correlation.

***Supporting Table 6.*** Observed case fatality rate by country/territory and by year.

***Supporting Figure 3.*** Annual predicted incidence by country/territory by year, 2001-2023.

***Supporting Figure 4.*** Comparison between observed and predicted cases.

***Supporting Figure 5.*** Observed case fatality rate between 1968 and 2022 and case fatality rate trend using Locally Estimated Scatterplot Smoothing (LOESS) model.

***Supporting Table 7.*** Annual estimated cases, case fatality rate and deaths and their 95% confidence interval, between 2001 and 2022.

